# Knowledge, attitude, practice and perceived barriers among healthcare professionals regarding COVID-19: A Cross-sectional survey from Pakistan

**DOI:** 10.1101/2020.04.13.20063198

**Authors:** Muhmmad Saqlain, Muhammad Muddasir Munir, Saif ur Rehman, Aqsa Gulzar, Sahar Naz, Zaheer Ahmed, Azhar Hussain Tahir, Muhammad Mashhood

**Affiliations:** Department of Pharmacy, Quaid-I-Azam University, Islamabad; Institute of Pharmaceutical sciences, University of Veterinary and Animal Sciences, 54000, Lahore; College of Physician and Surgeons, Pakistan; Institute of Pharmacy, Lahore College for Women University, Lahore; School of Pharmacy, The University of Faisalabad, Faisalabad; Department of Pharmacy, University of Sargodha, Islamabad; Department of Pharmacy, Government College University, Faisalabad

**Keywords:** KAP, COVID-19, healthcare workers, awareness

## Abstract

Coronavirus disease (COVID-19) is a highly transmittable infection and Pakistan faces sudden hike in number of positive cases including number of healthcare professionals (HCPs) also acquired infection. Knowledge, attitude, and practice survey provides a suitable format to evaluate existing programs and to identify effective strategies for behavior change in society. Therefore, the aim of study is to assess knowledge, attitude and practice among HCPs in Pakistan regarding COVID-19. An online survey-based study was conducted among healthcare professionals including physicians, pharmacists and nurses. A self-administered validated (Cronbach alpha= 0.077) questionnaire comprised of five sections (Demographics, Knowledge, attitude, practice and perceived barriers) were used for data collection. Of 414 participants, 29.98% (n=120) physicians, 46.65% (n= 189) pharmacists and 25.36% (n= 105%) nurses. Most commonly utilized information source was social media. Findings showed HCPs have good knowledge (93.2%, n=386), positive attitude (8.43±1.78) and good practice (88.7%, n=367) regarding COVID-19. HCPs perceived that overcrowding in emergency room (52.9%, n=219), limited infection control material (50.7%, n=210) and poor knowledge regarding transmission (40.6%, n=168) of COVID-19 are the major barriers in infection control practice. Binary logistic regression analysis demonstrated that HCPs of age group 40-49 years (OR: 1.419, 95%CI: (0.14-4.78, P=0.041) have higher odds of good knowledge. Similarly, age group of 31-39 years (OR: 1.377, 95% CI: 0.14-2.04, P=0.05), experience of more than 5 years (OR: 10.71, 95% CI: 2.83-40.75, P<0.001), and pharmacist job (OR: 2.247, 95% CI: 1.11-4.55, P=0.025) were the substantial determinants of good practice regarding COVID-19. HCPs in Pakistan have good knowledge, yet, there are areas where gaps in knowledge and practice was observed. To effectively control infection spread, well-structured training programs must be launched by government targeting all kinds of HCPs to raise their existed knowledge.

## 1. Introduction

In earlier December, first case of pneumonia of unknown cause originated in Wuhan, capital city of Province Hubei, China, and on 31 December 2019, with emergence of more such cases, Wuhan gained attention by World Health Organization (WHO) [1]. The pathogen identified was named as novel coronavirus (2019-nCoV), currently called as severe acute respiratory syndrome corona virus-2 (SARS-CoV-2), an enveloped and single stranded RNA virus [2] which has phylogenetic resemblance to SARS-COV-1 [3]. Owing to rapid spread of this deadly virus from epicenter to number of countries, WHO declared it as public health emergency of international concern (PHEIC) on January 30, 2020. Later, due to uncased fast spread, severity of illness, the continual escalation in number of affected countries, cases and causalities, WHO declared coronavirus disease 2019 (COVID-19) a global pandemic on 11 March 2020 [4]. To date (12 April, 2020), the COVID-19 have spread to 210 countries and territories accounted for 1,790,550 laboratory confirmed cases and 109,654 mortalities also attributed to this deadly pathogen [5].

Pakistan, a country with most vulnerable geographical location for this pandemic as it sandwiches between China; a country of origin and also a first epicenter of COVID-19 and Iran; an epicenter of Islamic world with 4,357 deaths and 70,029 cases attributed to COVID-19 [5]. Pakistan’s first COVID-19 case was reported on 26 February 2020, has travelled from Iran [6]. By 12 April 2020, Pakistan has 5,038 laboratory confirmed cases and 86 COVID-19 associated deaths. Punjab (n=2425) has highest number of cases followed by Sindh (n=1318), Khyber Pakhtunkhwa (KP) (n=696), Baluchistan (n=228), Gilgit Baltistan (216), Islamabad (n=119), and Azad Jammu Kashmir (n=35) [7].

COVID-19 transmits from person to person by droplets when an infected person sneezes and by direct contact and virus has an incubation period of 4-14 days [8]. Elderly and patients who suffered with chronic medical conditions like diabetes and cardiovascular diseases are more likely to get severe infection [8]. The main manifestations of COVID-19 are fever, dry cough, dyspnea, myalgia, fatigue, hypolymphaemia, and radiographic evidence of pneumonia [1,9]. Presently, no antiviral therapy or vaccine is explicitly recommended for COVID-19 and implementation of preventive measures to control COVID-19 is the mainstay critical intervention [10].

Healthcare professionals (HCPs) of all levels and kinds are primarily involved in catering patients of this highly transmittable pathogen. COVID-19 has posed serious occupational health risk to the HCP owing to their frequent exposure to infected individuals [11]. Protection of HCPs and prevention of intra-hospital transmission of infection are important aspects in epidemic response and this requires that HCPs must have updated knowledge regarding source, transmission, symptoms and preventive measures [12]. Literature suggest that lack of knowledge and misunderstandings among HCPs leads to delayed diagnosis, spread of disease and poor infection control practice [13]. Several thousand healthcare workers have already been infected, mainly in China [11]. In Pakistan, number of HCPs (15 medics in Multan, 17 doctors & 5 paramedics in Quetta, 16 medics in KP, 4 doctors & 1 paramedics in Karachi) are infected of COVID-19 [14,15] while unfortunately three doctors [16] and 1 nurse was died of COVID-19 [17]. Preventing intrahospital transmission of the communicable disease is therefore a priority.

Amidst to current pandemic, WHO has issued several guidelines and also started online courses and training sessions to raise awareness and preparedness regarding prevention and control of COVID-19 among HCPs [18]. In addition to WHO, National institute of Health (NIH), Islamabad, Pakistan also published several recommendations for HCPs aimed to reduce occupational spread of infection among HCPs [19]. Although educational campaigns have increased their awareness regarding COVID-19 yet it remains unclear to what extent this knowledge can be put into practice and to what extent this practice actually reduces COVID-19 infection spread. Knowledge, attitude, and practice survey provides a suitable format to evaluate existing programs and to identify effective strategies for behavior change in society [20]. Currently, there is scarce information regarding the awareness level of HCPs in Pakistan,

Therefore, the present study aimed to identify the current status of knowledge, attitude and practices regarding COVID-19 among healthcare professionals in Pakistan. In addition, study will highlight the information sources utilized and barriers in infection control perceived by HCPs.

## 2. Methods

### 2.1. Study design

A cross-sectional survey-based study was conducted during the month of March 2020, days of strict lockdown to implement social distancing to avoid spread of pandemic. As it was not feasible to conduct population-based survey in this critical condition, the investigators selected an online data collection method.

### 2.2. Sampling, study population and data collection method

Sample size calculated by Raosoft was 377 assuming a response rate of 50%, confidence interval (CI) 95%, Z as 1.96, and margin of error d as 5%. Considering, an additional 10% (n=37) for any error in questionnaire filling, a final sample size of 414 will be required. Survey was started on 25, March 2020, and response acceptance was closed (05-April 2020) when required sample size was achieved.

The study population eligible for participation in this survey were HCPs including doctors, pharmacists, and nurses. Pakistani nationals of age 16 years or more and consented for filling the data form are invited to participate.

A questionnaire was designed on google forms and link generated was shared on WhatsApp grou2ps of HCPs. Link was also shared personally to HCPs who were in contact list of investigators. Respondents from other provinces were also eligible to participate if they are willing to fill the questionnaire.

### 2.3. Measure

A survey instrument was designed based on extensive literature review, course material regarding emerging respiratory diseases including COVID-19 by WHO [18], and guidelines issued by NIH, Islamabad Pakistan [19]. After an initial draft of the questionnaire designed, it was validated in 2 steps. Firstly, the study instrument was sent to researchers and professionals from pharmacy and medical background to give their expert opinion with respect to its simplicity, relativity and importance. Secondly, a pilot study was conducted by selecting a small sample of health care professionals (n = 40) who gave their opinions on making the questionnaire simpler and shorter. Participants from all healthcare professions were selected for the pilot study. Amendments from the participants were considered and integrated into the questionnaire, while ensuring its consistency with the published literature. After a thorough discussion, questionnaire was finalized by the authors and subsequently distributed to the participants for their response. Reliability coefficient was calculated by using SPSS v.20 and the value of Cronbach’s alpha was found to be 0.77. The data of the pilot study was not used for the final analysis.

The questionnaire was consisted of questions assessing demographics, information source, knowledge, attitude, practice toward COVID-19 and perceived barriers in infection control practice (Supplementary file 1). Demographic characteristics included were gender, age, profession and experience, and one item regarding source of information about COVID-19.

Knowledge section comprised of 14 items and each question was responded as Yes, No and I don’t know. The correct answer was marked as 1 while wrong answer was marked as 0. Total score ranges from 0-14 and a cut off level of ≤10 was set for poor knowledge and ≥11 for good knowledge.

Attitude section comprised of 7 items and response of each item was recorded on 5-point Likert scale as follows strongly agree (1-point), agree (2-point), Undecided (3-point), disagree (4-point), and strongly disagree (5-point). Total score ranges from 7 to 35, with an overall lower mean score indicates positive attitude toward COVID-19.

Practice section included 6 items and each item was responded as Yes (1-point), No (0-point), and Sometimes (0-point). Practice items total score ranged as 0-6, and a score of 5-6 demonstrated good practice and a score of 1-4 indicates poor practice toward precautionary measures of COVID-19.

Seven items assessed the perception of HCPs regrading barriers in infection control practice. Each question was respondent on 5-point Likert scale as follows strongly agree, agree, undecided, disagree, and strongly disagree. Responses were presented as frequencies and percentages.

### 2.4. Ethics

The study was performed in accordance to declaration of Helsinki. Due to lockdown, universities were closed, hence study protocol was approved from Hospital board (756/THQ/HR). Study questionnaire contained consent portion that stated purpose, nature of survey, study objectives, volunteer participation, declaration of confidentiality and anonymity.

### 2.5. Statistical analysis

Data was entered in Microsoft Excel and later imported in SPSS V.21 for statistical analysis. Numerical variables were measured as mean and standard deviations while categorical variables were expressed as frequencies and percentages. Inferential statistics were applied depending upon nature of data and variables. Chi-square tests were applied to find difference in knowledge groups (good vs poor) and practice (good vs poor) by demographic characteristics. Independent sample t-test and one-way ANOVA analysis were performed in assessing any difference in mean attitude score by demographic characteristics. Pearson-rank correlation tests were applied to find any correlation between knowledge, attitude and practice sections. To find possible determinants of good knowledge and practice, a binary logistic regression analysis will be applied and expressed as odds ratio (OR) and 95% confidence interval (CI). A p value of less than 0.05 will be considered as significant in all tests.

## 3. Results

## 3.1. Characteristics of HCPs

A total of 414 respondents were included in final analysis, of which 29.98% (n=120) physicians, 46.65% (n= 189) pharmacists and 25.36% (n= 105) nurses. There was equal proportion of male (50.5%, n=209) and female (49.5%, n=205) respondents, majority (74.9%, n=310) of respondents were of age less than 30 years, and 31.6% (n=131) have experience of 1-3 years (**Table I**).

**Table I.**
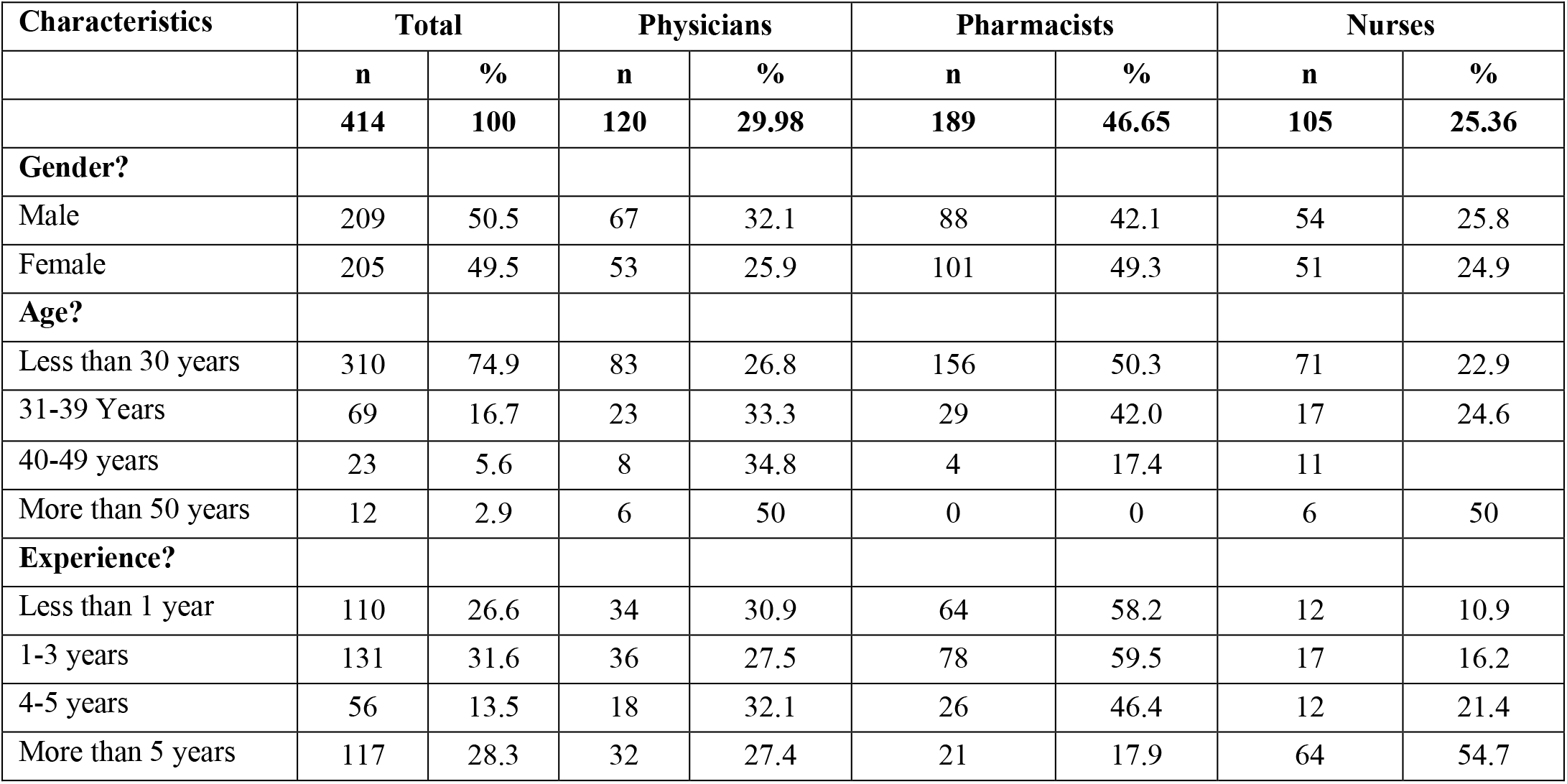
Demographics of Healthcare professionals (N=414).

Figure 1. summarizes the sources of information utilized by HCPs to seek information regarding COVID-19. Majority of professionals reported social media (87.68%, n=363) as main source of information followed by radio & television (45.89%, n=190) and seniors/other colleagues (42.51%, n=176) (Fig.1).

**Figure 1.**
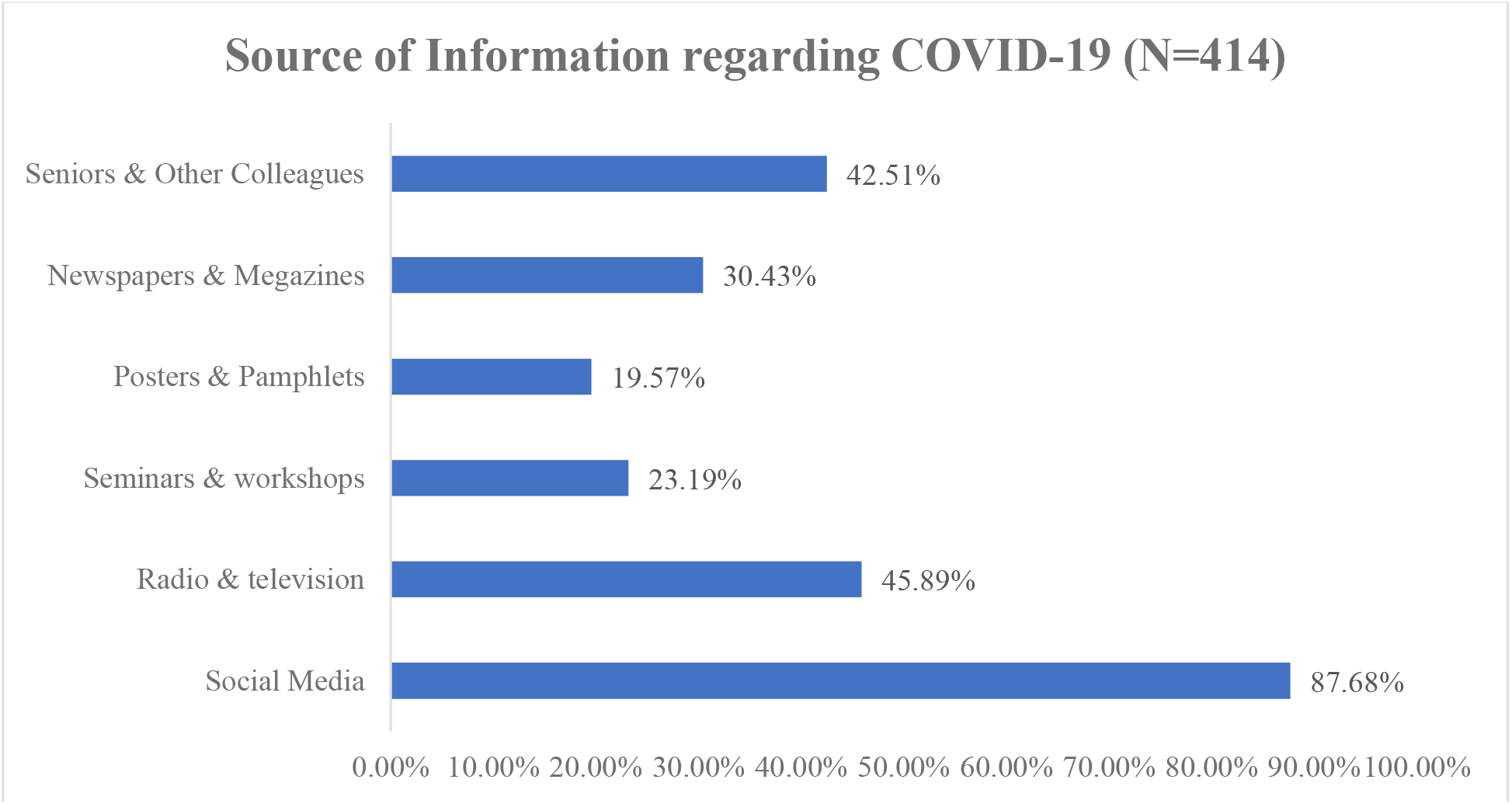
Information sources reported by healthcare professionals.

### 3.2. Knowledge among HCPs regarding COVID-19

Fig. 2 represents the responses obtained for knowledge items of questionnaire. Mixed responses were obtained regarding 14 knowledge items. All the respondents correctly answered that COVID-19 is a viral infection. More than 90% of HCPs were well aware about fatality of disease, incubation period, symptoms, vaccination availability status, transmission and precautions regrading COVID-19. Additionally, 82.13% (n==340) correctly identified that antibiotics are not first line treatment, and 87.92% (n=364) respondents were also well aware of that plant is not a source of infection. When questions asked regarding risky groups, testing, and influenza vaccine protection, 21.01%, 23.91% and 23.19% respondents respectively were unable to identify correct responses (Fig. 2).

**Figure 2.**
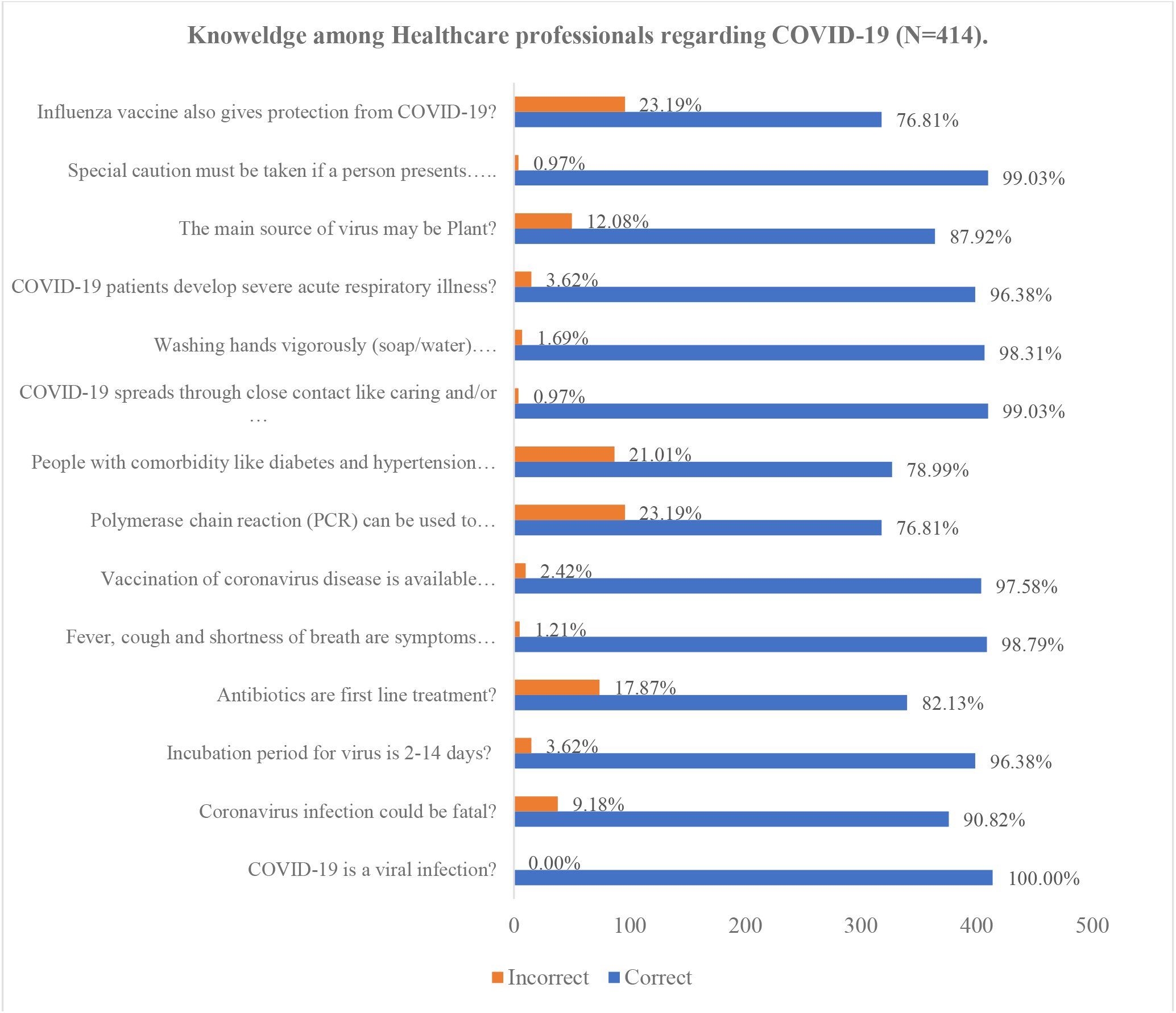
Knowledge of healthcare professionals regarding COVID-19.

### 3.3. Attitude among HCPs regarding COVID-19

Fig. 3 represents the responses obtained for attitude items. All the HCPs responded to all 7 items on their attitude regarding COVID-19. Findings demonstrated highly positive attitude of HCPs towards COVID-19. About 97.80% (n=405) individuals strongly agreed that gowns, gloves must be used when dealing with infected patients and equal proportion of respondents strongly believed that diseased patients should be kept isolated. More than 80% participants strongly agreed that transmission of COVID-19 could be prevented by following universal precautions given by WHO, CDC and that healthcare workers must acknowledge themselves with all the information regarding COVID-19. Similarly, 76.80% (n=318) HCPs had positive attitude regarding “intensive and emergency treatment should be given to diagnosed patients”, and 70% participants strongly agreed that “any related information should be disseminated among healthcare workers”. On the other hand, 69.10% (n=286) HCPs strongly agreed that prevalence of infection could be controlled by actively involving in infection control programs (Fig.3).

**Figure 3.**
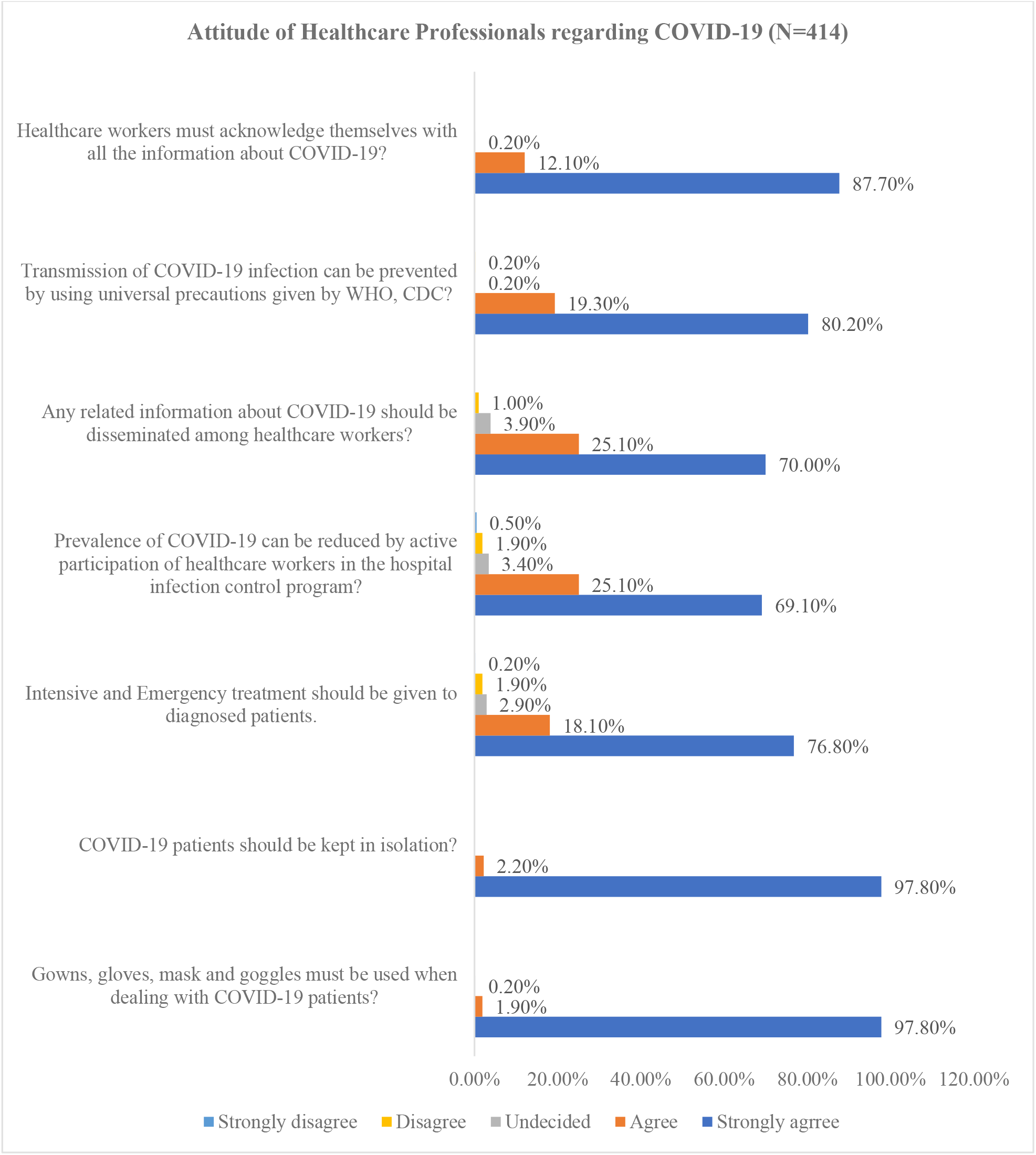
Attitude of healthcare professionals regarding COVID-19.

### 3.4. Practice among HCPs toward COVID-19

Fig. 4 represents the responses obtained for practice assessing items of questionnaire. Majority of respondents had good practice regrading each item with highest practice showed among HCPs towards washing of hands with soap or cleaning with sanitizers (96.10%, n=398). A lower percentage of good practice was observed among HCPs in avoiding to touching of eyes, nose or mouth (84.30%, n=349) (Fig.4).

**Figure 4.**
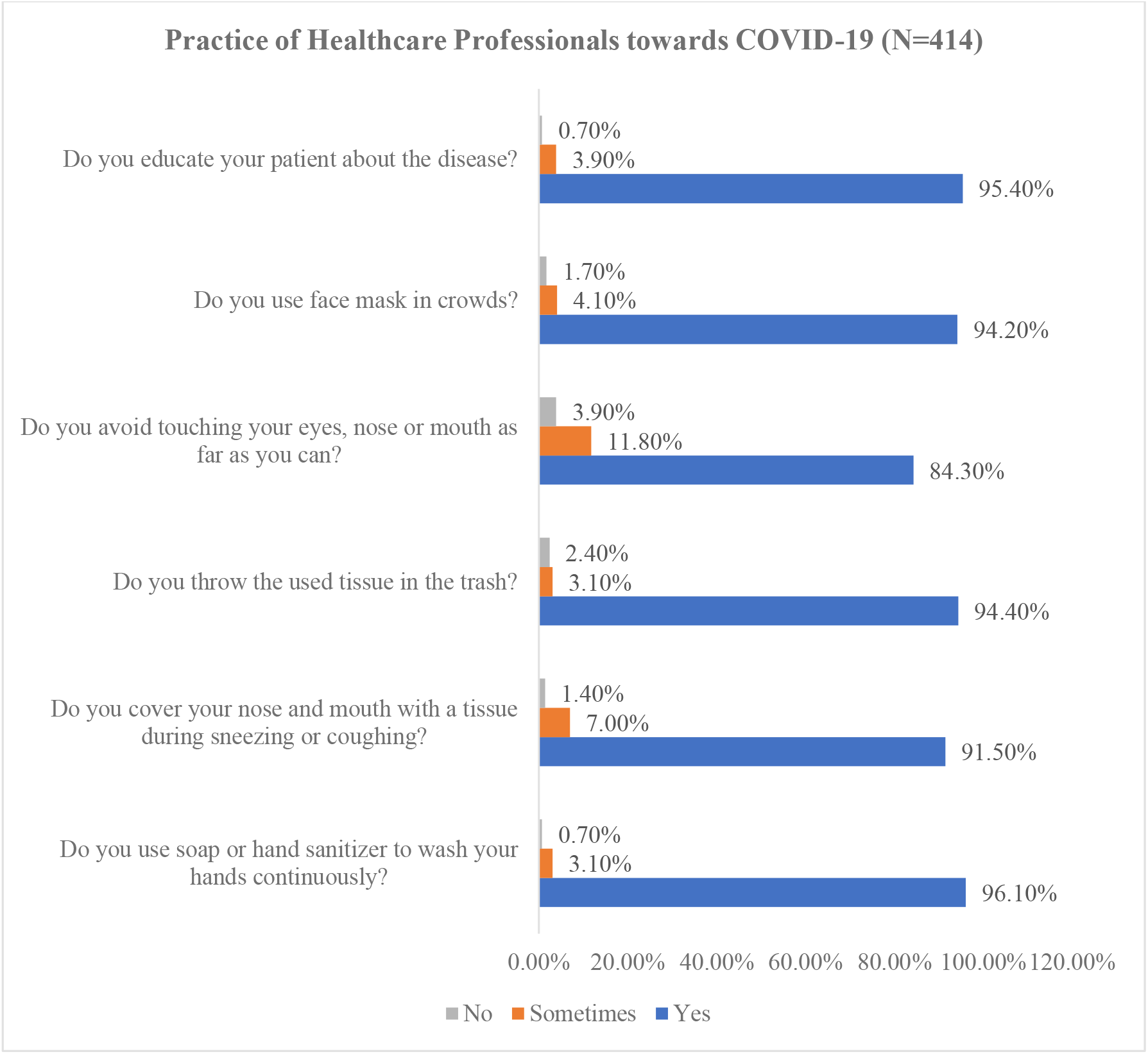
Practice among healthcare professionals regarding COVID-19.

### 3.5. Barriers perceived by HCPs in infection control practice

Mixed perception was reported by HCPs regarding barriers in infection control practice. Of 414 participants, 40.6% (n=168) professionals presumed that lack of knowledge about transmission of virus is a barrier, and 50.7% (n=210) strongly agreed that limited infection control material is also a barrier towards control of infection. Overcrowding in emergency room was perceived as barrier by majority (52.9%, n=219) of HCPs. On the other hand, 31.6% (n=131) and 36.7% (n=152) participants believed that not wearing a mask, and no hand washing is not any barrier in infection control (Fig. 5) (Fig.5).

**Figure 5.**
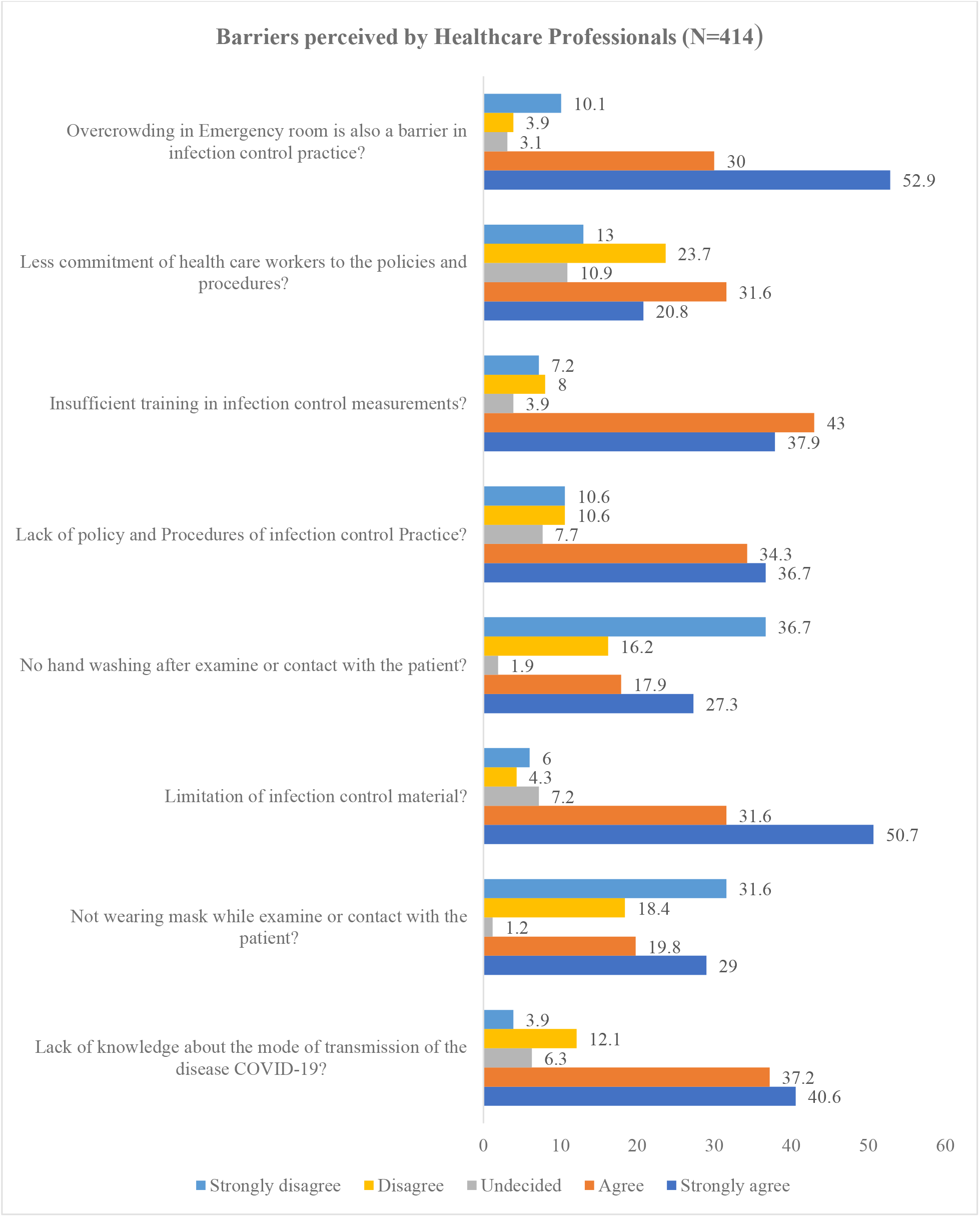
Barriers in infection control practice perceived by healthcare professionals regarding COVID-19.

### 3.6. Difference in knowledge, attitude, and practice among HCPs regarding COVID-19

Majority of respondents (93.2%, n=386) had good knowledge regarding COVID-19. Chi-square tests were applied to find difference in knowledge status by demographic characteristics of HCPs towards COVID-19. Findings demonstrated that age was significantly associated with good knowledge as 95.5% (n=284) HCPs of age less than 30 years had good knowledge compared to healthcare professions of other age groups. The overall knowledge score indicated good knowledge among physicians (93.3%, n=112), pharmacists (94.7%, n=179), and nurses (90.5%, n=95) but the difference was statistically insignificant (P=0.383). Similarly, knowledge score was not statistically differed by gender (P=0.403), and experience (P=0.281) (**Table II**).

Independent sample t-test revealed that attitude score was not statistically (P=0.788) differed by gender (8.41 vs 8.45). One-way ANOVA analysis indicated that physician (8.41) have highest positive attitude followed by pharmacists (8.43) and nurses (8.64) but association was not significant (P=0.358). Similarly, no difference in mean attitude score was found by age and experience (P>0.05). (**Table II**)

**Table II.** Difference in Healthcare Professional’s Knowledge, Attitude and Practice by demographics (N=414)

| Characteristics | Knowledge* |  |  | Attitude** |  |  | Practice*** |  |  |
| --- | --- | --- | --- | --- | --- | --- | --- | --- | --- |
|  | Poor Knowledge | Good Knowledge | X<sup>2</sup> (P) | Mean | SD | t/F (P) | Poor Practice | Good Practice | X<sup>2</sup> (P) |
| **Overall** | **28 (6.8)** | **386 (93.2)** |  | **8.43** | **1.78** |  | **47 (11.3)** | **367 (88.7)** |  |
| **Gender?** |  |  | 0.699 (0.403) |  |  | 0.269 (0.788) |  |  | 0.286 (0.592) |
| Male | 12 (5.7) | 197 (94.3) |  | 8.41 | 1.77 |  | 22 (10.5) | 187 (89.5) |  |
| Female | 16 (7.8) | 189 (92.2) |  | 8.46 | 1.79 |  | 25 (12.2) | 180 (87.8) |  |
| **Age?** |  |  | 9.836 (0.020) |  |  | 0.198 (0.699) |  |  | 0.295 (0.957) |
| Less than 30 years | 26 (8.4) | 284 (91.6) |  | 8.44 | 1.73 |  | 36 (11.6) | 274 (88.4) |  |
| 31-39 Years | 5 (7.2) | 64 (92.8) |  | 9.43 | 2.07 |  | 8 (11.6) | 61 (88.4) |  |
| 40-49 years | 1 (4.5) | 22 (95.5) |  | 8.26 | 1.51 |  | 2 (8.7) | 21 (91.3) |  |
| More than 50 years | 2 (16.7) | 10 (93.2) |  | 8.16 | 1.65 |  | 1 (8.3) | 11 (91.7) |  |
| **Experience?** |  |  | 3.059 (0.281) |  |  | 1.347 (0.259) |  |  | 9.827 (0.012) |
| Less than 1 year | 6 (5.5) | 104 (94.5) |  | 8.78 | 1.99 |  | 19 (17.3) | 91 (82.7) |  |
| 1-3 years | 12 (9.2) | 119 (90.8) |  | 8.53 | 1.81 |  | 16 (12.2) | 115 (87.8) |  |
| 3-5 years | 5 (8.9) | 51 (91.1) |  | 8.34 | 1.69 |  | 7 (12.5) | 49 (87.5) |  |
| More than 5 years | 5 (4.3) | 112 (95.7) |  | 8.25 | 1.68 |  | 5 (4.3) | 112 (95.7) |  |
| **Profession?** |  |  | 1.920<br>(0.383) |  |  | 1.030<br>(0.358) |  |  | 4.802<br>(0.091) |
| Physician | 8 (6.7) | 112 (93.3) |  | 8.41 | 1.78 |  | 6 (5.7) | 114 (94.3) |  |
| Pharmacists | 10 (5.3) | 179 (94.7) |  | 8.43 | 1.71 |  | 18 (9.5) | 171 (90.5) |  |
| Nurses | 10 (6.8) | 95 (90.5) |  | 8.64 | 1.88 |  | 9 (8.6) | 96 (91.4) |  |

\* Knowledge section total score ranges from 0-14 and a cut off level of $\leq 10$ was set for poor knowledge and $\geq 11$ for good knowledge. \*\* Attitude section total score ranges from 7 to 35, with an overall lower mean score indicates positive attitude toward COVID-19. \*\*\* Practice items total score ranged as 0-6, and a score of 1-4 indicates poor practice regarding COVID-19.
**Note:** A p value of less than 0.05 considered significant. Bold values showing significant differences.

Of 414 participants, 88.7% (n=367) had positive practice (score=5-6) in following precautions to avoid COVID-19. Chi-square analysis revealed that majority (95.7%, n=112) of HCPs having greater experience (more than 5 years) had good practice of following precautions compared to less experienced counterparts (P=0.020). Findings also showed that physician (94.3%, n=114) had highest practice than nurses (91.4%, n=96), and pharmacists (90.5%, n=171), but the difference was not significant (0.091). (**Table II**).

### 3.7. Logistic regression analysis for factors associated with Good knowledge and Practice regarding COVID-19

Binary logistic regression analysis demonstrated that age group of 40-49 years (OR: 1.419, 95%CI: 0.14-4.78, P=0.041) and age group of 50 years or more were the significant factors associated with good knowledge. Similarly, age group of 31-39 years (OR: 1.377, 95% CI: 0.14-2.04, P=0.05), experience of more than 5 years (OR: 10.71, 95% CI: 2.83-40.75, P<0.001), and pharmacist job (OR: 2.247, 95% CI: 1.11-4.55, P=0.025) were the significant determinants of good practice regarding COVID-19 (**Table III**).

**Table III.** Logistic regression analysis for factors associated with Good knowledge and Practice regarding COVID-19 (N=414).

| Characteristics | Knowledge* |  |  |  | Practice** |  |  |  |
| --- | --- | --- | --- | --- | --- | --- | --- | --- |
|  | Poor Knowledge | Good Knowledge | OR | P | Poor Practice | Good Practice | OR | P |
|  | N (%) | N (%) | 95% CI |  | N (%) | N (%) | 95% CI |  |
| **Gender?** |  |  |  |  |  |  |  |  |
| Male | 12 (5.7) | 197 (94.3) | 1.00 | - | 22 (10.5) | 187 (89.5) | 1.00 | - |
| Female | 16 (7.8) | 189 (92.2) | 0.696<br>(0.301-1.608) | 0.396 | 25 (12.2) | 180 (87.8) | 0.977<br>(0.51-1.89) | 0.445 |
| **Age?** |  |  |  |  |  |  |  |  |
| Less than 30 years | 26 (8.4) | 284 (91.6) | 1.00 | - | 36 (11.6) | 274 (88.4) | 1.00 | - |
| 31-39 Years | 5 (7.2) | 64 (92.8) | 1.398<br>(0.06-2.63) | 0.089 | 8 (11.6) | 61 (88.4) | 1.377<br>(0.14-2.04) | **0.05** |
| 40-49 years | 1 (4.5) | 22 (95.5) | 1.419<br>(0.14-4.78) | **0.041** | 2 (8.7) | 21 (91.3) | 1.406<br>(0.07-2.93) | 0.313 |
| More than 50 years | 2 (16.7) | 10 (93.2) | 1.497<br>(0.17-4.18) | **0.038** | 1 (8.3) | 11 (91.7) | 1.444<br>(0.04-4.73) | 0.501 |
| **Experience?** |  |  |  |  |  |  |  |  |
| Less than 1 year | 6 (5.5) | 104 (94.5) | 1.00 | - | 19 (17.3) | 91 (82.7) | 1.00 | - |
| 1-3 years | 12 (9.2) | 119 (90.8) | 1.555<br>(1.19-2.54) | 0.260 | 16 (12.2) | 115 (87.8) | 1.453<br>(0.69-3.03) | 0.319 |
| 3-5 years | 5 (8.9) | 51 (91.1) | 1.619<br>(1.26-2.84) | 0.190 | 7 (12.5) | 49 (87.5) | 2.11<br>(0.73-6.11) | 0.169 |
| More than 5 years | 5 (4.3) | 112 (95.7) | 1.999<br>(1.29-4.81) | 0.481 | 5 (4.3) | 112 (95.7) | 10.71<br>(2.83-40.75) | **<0.001** |
| **Profession?** |  |  |  |  |  |  |  |  |
| Physician | 8 (6.7) | 112 (93.3) | 1.00 | - | 6 (5.7) | 114 (94.3) | 1.00 | - |
| Pharmacists | 10 (5.3) | 179 (94.7) | 1.44<br>(0.54-3.87) | 0.461 | 18 (9.5) | 171<br>(90.5) | 2.247<br>(1.11-4.55) | **0.025** |
| Nurses | 10 (6.8) | 95 (90.5) | 0.616<br>(0.21-1.78) | 0.369 | 9 (8.6) | 96 (91.4) | 1.178<br>(0.48-2.90) | 0.724 |

OR: Odds ratio, CI: Confidence interval.
\* Knowledge section total score ranges from 0-14 and a cut off level of $\leq 10$ was set for poor knowledge and $\geq 11$ for good knowledge. \*\*\* Practice items total score ranged as 0-6, and a score of 1-4 indicates poor practice regarding COVID-19. Note: A p value of less than 0.05 considered significant. Bold values shoeing significant differences.

Pearson correlation tests revealed a statistically significant positive linear correlation between knowledge, attitude and practice scores as follows: knowledge-attitude (r = 0.106, p value = .030), knowledge-practice (r = 0.142, p value = .016), and attitude-practice (r = 0.174, p value = .004) (**Table IV**).

**Table IV.** Correlation between scores of knowledge, attitude, and practice

| Variable | Correlation Coefficient | P-Value |
| --- | --- | --- |
| Knowledge-Attitude | 0.106* | .030 |
| Attitude-Practice | 0.174* | .004 |
| Knowledge-Practice | 0.142* | .016 |

\* Correlation significant at 0.05 level (2 tailed).

## 4. Discussion

To best of our knowledge, this is the first study that has thoroughly assessed the knowledge, attitude and practice of HCPs toward COVID-19 in Pakistan. The study also highlighted possible barriers perceived by HCPs in infection control practices.

For HCPs, good knowledge, positive attitude, and good practices of following precautionary measures such as wearing gloves, protective clothing, goggles and face mask is imperative in effective dealing with infected patients with minimum risks. Also, ongoing pandemic nature of disease made it necessary for HCPs to multiply their alarms corresponding to critical situation and to put efforts in following and implementing related hygienic conditions as well as recommendations. On the other hand, healthcare professional’s good knowledge and practice in complying precautionary measures creates awareness among patients as well as gives an important message in society [21].

Findings of current survey demonstrated that majority of HCPs have good knowledge (93.2%, n=386), positive attitude (mean 8.43) and good practice (88.7%, n=367) towards COVID-19. Of note that, 87.68% HCPs utilized social media as a main source of information while only 23.19% seek information from seminars & workshops. Findings are consistent with number of studies as Giao et al reported social media as a main source of information regarding COVID-19 used by 91.1% participant health care workers [22]. Similarly, a study conducted among 453 HCPs found that more than 60% respondents use social media to seek information regarding COVID-19 [10].

This is important as in this global pandemic, there is also a pandemic of misinformation regarding COVID-19; a serious concern might lead to xenophobia in the world as already warned by scientists and WHO officials [23][24]. Also, there is plethora of malicious and unverified information waved on internet that spread quickly and could misguide the HCPs. So, situation demands that HCPs should carefully evaluate COVID-19 information sources and utilize authentic and valid content to seek information [10].

Current findings of good knowledge among HCPs is in line with findings of Giao et al who reported that 88.4% participants have sufficient knowledge regarding COVID-19 [22]. Shi et al also reported that 89.51% healthcare workers have claimed good knowledge while in another study conducted among nurses reported that 56.5% respondents have sufficient knowledge regarding transmission, symptoms and treatment of COVID-19 [12]. Present findings provide confidence in terms of healthcare professional’s knowledge regarding COVID-19 symptoms, transmission and preventive measures. This is of more significance in current scenario when there is no vaccine and research is ongoing so HCPs must aware of all the updates and take precautions in treating and preventing the infection.

Of note that, more than 20% health care professionals had inaccurate knowledge that influenza vaccine provide protection against COVID-19 and also were not aware of that patients with diabetes and hypertension are more likely to get infections. Despite of a prioritized global health emergency and availability of easily accessible sources provided by both national (National Institute of Health (NIH, Islamabad)) and international (WHO) healthcare authorities, these findings have shown knowledge gap among HCPs. This proportion (20%) is higher than other study that stated only 9.1% respondents have answered that flu vaccine could provide protection against COVID-19 [10]. Possible speculation is that currently the outbreak of COVID-19 in Pakistan is recent and discussion happens among HCPs is more likely regarding the symptoms of disease compared to other aspects like risky population and usefulness of existed vaccines and treatments. It is therefore necessary that these aspects of COVID-19 should be uncovered and cleared to HCPs so that they can educate people to counter this global public health issue.

Interestingly, pharmacists have higher knowledge than physicians and nurses but results were insignificant (P=0.383). Giao et al also found that pharmacists have higher knowledge compared to other HCPs regarding COVID-19 (P=0.015). Khan et al and Albarrak et al also found that allied health care workers including pharmacists have higher knowledge towards MERS compared to physicians [25,26]. While another study by Bhagavathula et al reported that physicians have higher knowledge compared to pharmacists [10]. Possible speculation could be that there are disparities of knowledge among HCPs. In addition to doctors, pharmacists also actively involved in seeking information owing to their active role in improving treatment outcomes of COVID-19 patients. Also, findings encourage a collaborative approach among different HCPs in relation to clinical decision making.

Findings showed highly positive attitude among all the HCPs towards wearing gowns and in following other recommendations. Possible explanation is that good knowledge among HCPs might lead to positive attitude towards COVID-19. This is also augmented by positive linear correlation between knowledge and attitude found in present study. Giao et al and Bhagavathula et al also reported that majority of HCPs have positive attitude towards COVID-19 [10,22].

Interestingly, participant’s attitude didn’t differ statistically (P>0.05) by age, gender, experience and job. Giao et al also found that attitude level regarding COVID-19 didn’t have any association with age (P=0.151), gender (P=0.129), and experience (P=0.453) but found statistically significant association with occupation/job [22]. While Albarrak et al and khan et al didn’t found any difference in attitude towards MERS among doctors, pharmacists, and nurses (P>0.05) [25,26].

Results revealed that majority of HCPs have good practice in following precautionary measures. Highest (96.10%) good practice was observed in washing hands with soap which is similar to the findings of Nour et al and khan et al who reported 95.4% and 85.7% HCPs used to wash their hands continuously [25,27]. Findings indicated pharmacist have higher odds of showing good practice compared to other HCPs. Possible explanation is that pharmacist is an emerging profession especially in developing countries like Pakistan, and they are actively involved in educational and training activities to seek knowledge and made themselves competent enough to involve in clinical decision making with other HCPs [28,29]. Also, pharmacists are actively involved in counselling the patients which leads to positivity in their attitude and good practice. Regression analysis also indicated that experienced health care professionals have good practice in following guidelines recommendations especially wearing of face mask as it is evident that use of personal protective equipment might be helpful in reducing spread of virus in hospital and protect others from infection [30]. Therefore, it is important among different workers to adhere to the practice guidelines according to NIH and WHO intending the COVID-19 infection in the community. The study further recommends healthcare workers to reinforce their knowledge and attitude that will eventually translate into good practice.

Findings demonstrated that majority of HCPs perceived that overcrowding of emergency department, limited infection control material and lack of knowledge regarding transmission of COVID-19 are the major barriers in infection control practice. These findings are important and should be addressed by Government and policy makers to establish effective policies focusing aforementioned barriers to control infection and ultimately spread of disease.

Correlational analysis also augments the findings of present study as knowledge is significantly related to attitude and also reflected by good practices. This view also inferred that HCPs with positive attitude are more interested in seeking knowledge and then put knowledge into practice. This correlation could also be explained by Reasoned action theory. This theory states that a person’s intention to a specific behaviour is a function of their attitude towards that behaviour [31]. Future studies should be conducted to understand possible factors that underlies knowledge pattern and attitudes expressed by HCPs.

## Strengths and Limitations

Current study highlighted the less explored area where scarce literature was available. Study identify the current status of HCPs knowledge; an important aspect in successful response to any epidemic. Questionnaire was developed by using WHO published material and a two-step validation approach also increases the reliability of current analysis.

The study has number of implicit limitations. Firstly, it is a cross-sectional study conducted during lockdown period, and universities were also closed, therefore institutional review board was not approached. Secondly, this is an online survey, responses mainly depend upon honesty and partly affected by recall ability and thus may subject to recall bias. Potential sample clustering might also limit the generalizability of study.

## Conclusion

Findings providing confidence as HCPs have good knowledge, positive attitude and good practice regarding COVID-19. The study also able to highlight gaps in specific aspects of knowledge, and practice that should be focused in future awareness and educational campaigns. Findings also demonstrated that HCPs were using less authentic sources, this aspect should immediately be addressed as it ultimately affects knowledge and reflected in attitude and practice. The study recommends the ministry of health authorities to promote all precautionary and preventive measures of COVID-19 with a comprehensive training program consisting better structured targeting all HCPs including physicians, pharmacists and nurses, in order to have equilibrium clinical knowledge about COVID-19.

## Data Availability

The data sets used or analyzed during the current study available from the corresponding author upon reasonable request.

## Acknowledgements

The authors would like to extend heartfelt graciousness to all the participants and teachers who provided support at every step of the research.

## Authors’ contributions

The manuscript idea, concept, writing, and layout was done by MS, MM and AG. MMM and SN provided critical help in writing, statistical and layout designing. SUR and ZA provided critical input regarding data analysis at every step of the manuscript writing process. MS, SUR and ZA proof read the manuscript and provided input in formulating the draft.

## Funding

The authors declare that they have not received any direct or indirect funding from any organization.

## Declaration of Interests

The authors have no relevant affiliations or financial involvement with any organization or entity with a financial interest in or financial conflict with the subject matter or materials discussed in the manuscript. This includes employment, consultancies, honoraria, stock ownership or options, expert testimony, grants or patents received or pending, or royalties.

## Notes

### Competing Interest Statement

The authors have declared no competing interest.

### Funding Statement

None of the authors have received any funding from any person or any organization.

